# Analysis of the epidemic growth of the early 2019-nCoV outbreak using internationally confirmed cases

**DOI:** 10.1101/2020.02.06.20020941

**Authors:** Qingyuan Zhao, Yang Chen, Dylan S Small

## Abstract

**Background:** On January 23, 2020, a quarantine was imposed on travel in and out of Wuhan, where the 2019 novel coronavirus (2019-nCoV) outbreak originated from. Previous analyses estimated the basic epidemiological parameters using symptom onset dates of the confirmed cases in Wuhan and outside China.

**Methods:** We obtained information on the 46 coronavirus cases who traveled from Wuhan before January 23 and have been subsequently confirmed in Hong Kong, Japan, Korea, Macau, Singapore, and Taiwan as of February 5, 2020. Most cases have detailed travel history and disease progress. Compared to previous analyses, an important distinction is that we used this data to informatively simulate the infection time of each case using the symptom onset time, previously reported incubation interval, and travel history. We then fitted a simple exponential growth model with adjustment for the January 23 travel ban to the distribution of the simulated infection time. We used a Bayesian analysis with diffuse priors to quantify the uncertainty of the estimated epidemiological parameters. We performed sensitivity analysis to different choices of incubation interval and the hyperparameters in the prior specification.

**Results:** We found that our model provides good fit to the distribution of the infection time. Assuming the travel rate to the selected countries and regions is constant over the study period, we found that the epidemic was doubling in size every 2.9 days (95% credible interval [CrI], 2 days—4.1 days). Using previously reported serial interval for 2019-nCoV, the estimated basic reproduction number is 5.7 (95% CrI, 3.4—9.2). The estimates did not change substantially if we assumed the travel rate doubled in the last 3 days before January 23, when we used previously reported incubation interval for severe acute respiratory syndrome (SARS), or when we changed the hyperparameters in our prior specification.

**Conclusions:** Our estimated epidemiological parameters are higher than an earlier report using confirmed cases in Wuhan. This indicates the 2019-nCoV could have been spreading faster than previous estimates.

## Main text

On December 31, 2019, the Health Commission in Wuhan, China, announced 27 cases of viral pneumonia. On January 8, 2020, the Chinese Center for Disease Control and Prevention announced that a novel coronavirus was the causative pathogen and was subsequently designated as 2019-nCoV. Anxiety quickly grew in the general public after a preeminent Chinese epidemiologist, Dr Zhong Nanshan, confirmed human-to-human transmission in a televised interview in the evening of January 20, and three days later the entire city of Wuhan was put into quarantine: all public transportation was suspended, and outbound trains and flights were halted. As of February 6, 2020, more than 28,000 cases of 2019-nCoV have been confirmed globally.

Using the first 425 confirmed patients of 2019-nCoV in Wuhan, a previous report estimated that the epidemic doubled in size every 7.4 days. With a mean serial interval of 7.5 days, the basic reproduction number was estimated to be 2.2 (95% confidence interval [CI], 1.4 – 3.9) (Li et al., 2020). Using internationally exported cases from Wuhan (known days of symptom onset from December 25, 2019 to January 19, 2020) and flight travel data, another report estimated that the basic reproduction number is 2.7 (95% CrI, 2.5 – 2.9) (Wu et al., 2020). Other analyses using the series of new confirmed cases have reported larger estimates of the basic reproduction number (Read et al., 2020; Zhao et al., 2020).

## Methods

### Dataset preparation and visualization

Using information published by governments and news media, we constructed a dataset of 2019-nCoV patients that are confirmed outside mainland China as of February 4, 2020. We only kept cases who resided in Wuhan or had recent travel history to Wuhan. We excluded patients who arrived in these countries and regions after January 23 or were reported to have exposure to 2019-nCoV outside Wuhan. For each cluster of cases (two cases are considered to belong to the same cluster if they had close contact outside Wuhan), we kept the first confirmed case and discarded the other cases as they were likely infected during their travel outside Wuhan. We only considered cases confirmed in Hong Kong, Japan, Korea, Macau, Singapore, and Taiwan as these governments provided detailed information on the confirmed cases. After applying this selection criterion, we obtained a sample of 46 coronavirus cases who were most certainly exported from Wuhan.

Based on the trajectories of these cases, we simulated the dates they were infected as follows. Among the 46 cases, one (18^th^ case in Singapore) has missing symptom onset dates. We used the *mice* software package (van Buuren & Groothuis-Oudshoorn, 2011) to obtain 100 imputations of the missing symptom onset date. For each imputation of symptom onset, we simulated the infection time based on Gamma-distributed incubation period that are have the same mean (5.2 days) and 95th percentile (12.5 days) as previous estimates (Li et al., 2020). We repeated the simulation until the infection time is consistent with the case’s trajectory and known exposure to other confirmed cases. In particular, because infection must be prior to the patient’s arrival if we assume all the cases were infected in Wuhan, this allows us to greatly narrow down the infection time. We repeated the simulation of infection time for 100 times and obtained a distribution of infection time for the 46 cases (Figure 1).

**Figure 1.**
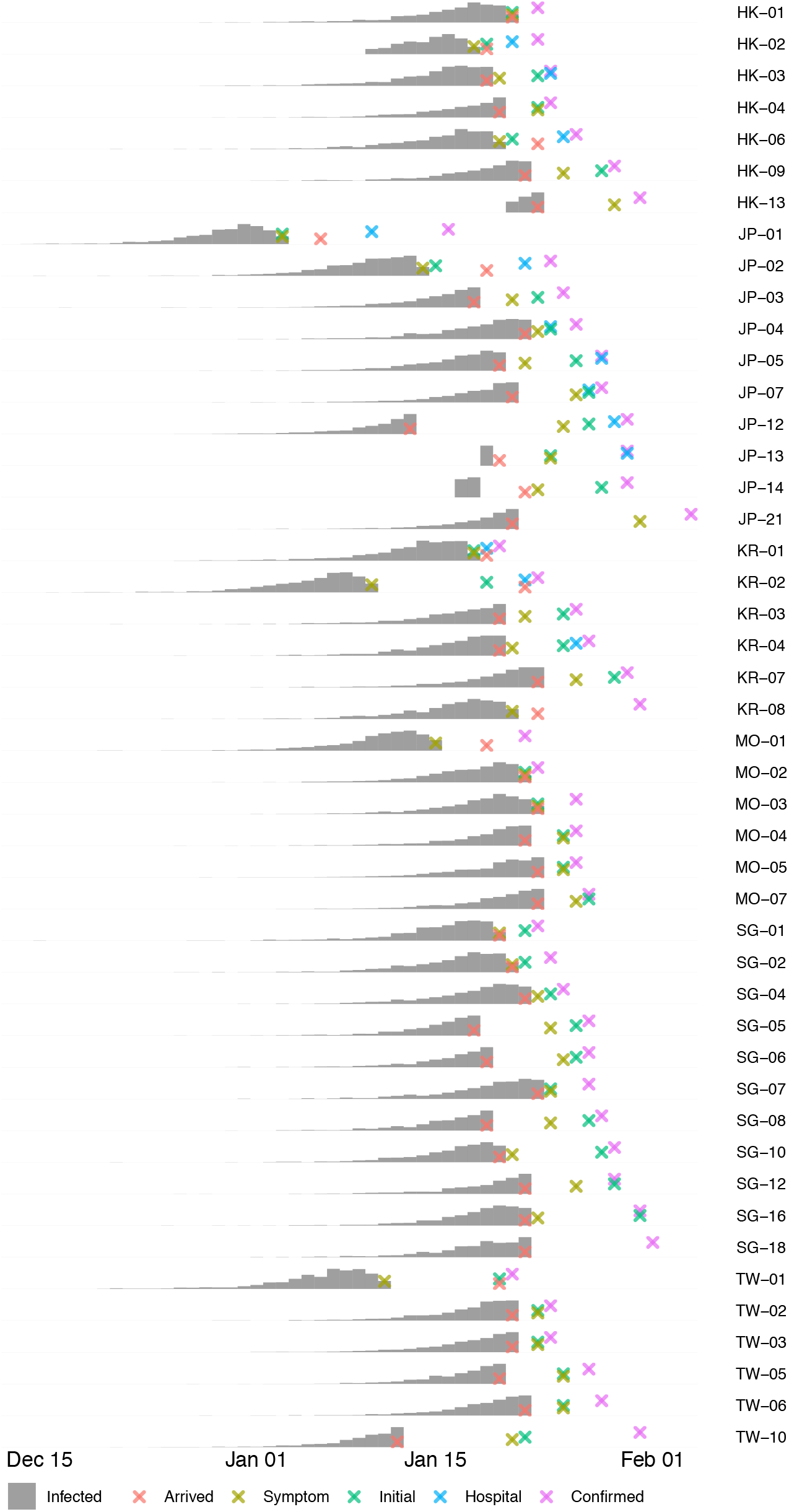
Trajectory and distribution of simulated infection time of 46 cases of 2019-nCoV confirmed in 6 Asian countries and regions (HK: Hong Kong, JP: Japan, KR: Korea, MO: Macau, SG: Singapore, TW: Taiwan). Infected: when the patient was infected; Symptom: when the patient first showed symptoms; Initial: when the patient first sought medical consultation or was isolated as a suspected case; Hospital: if the patient was not immediately admitted after the initial visit, when the patient was finally admitted to a hospital; Confirmed: when the case was confirmed as 2019-nCoV positive.

### Statistical model

We modeled, *WI*_*t*_, the number of new infections in Wuhan on day *t* since December 1, 2019 and assume it was growing exponentially up till January 23:

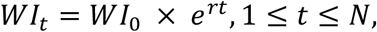

where *r* is the growth exponent and *N* = 31 + 23 = 54 is the length of the series considered. We assumed, *OI*_*t*_, the number of new infections in Wuhan on day *t* who subsequently traveled to one of the selected Asian countries and regions, follows a Poisson distribution:

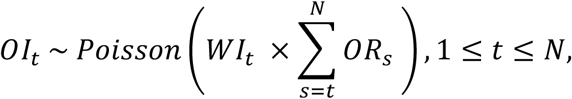

where *OR*_*s*_ is the rate of travel on day *s*. The sum is from *t* to *N* because each person infected on day *t* can only travel out of Wuhan from day *t* to day *N* (corresponding to January 23, 2020 when all outbound trains and flights were halted). Since we collected this dataset 13 days after the outbound travel e ban, which is more than the estimated 95^th^ percentile of the incubation period (Li et al., 2020), we assume that all 2019-nCoV cases traveled from Wuhan to the selected countries and regions have already been confirmed.

We expected that the international travel from Wuhan was relatively stable during this period. This motivated our working model for *OR*_*s*_:

1. *OR*_*s*_ = *OR* for the entire study period;

We considered another two choices of the travel rate as sensitivity analysis:

2. *OR*_*s*_ = *OR* before January 21 and *OR*_*s*_ = 2 × *OR* on and after January 21;
3. *OR*_*s*_ = 0 before January 23 and *OR*_*s*_ = *N* × *OR* on January 23.

The second choice is intended to model the possible panic effect after human-to-human transmission was publicly confirmed. The third choice is impossibly extreme. We only included it to compare our results with assuming a simple exponential model for *OI*_*t*_ with no offset, thereby ignoring the effect of the January 23 travel ban.

### Model fitting and statistical inference

We computed, *E*[*OI*_*t*_], the mean of *OI*_*t*_ using the simulated distribution of infection dates. Using data from January 1 to January 15, we fitted the following linear regression for log (*E*[*OI*_*t*_]) with offset log(*N* − *t* + 1) that is derived from the statistical model above assuming constant travel rate:

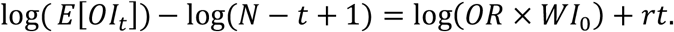

We compared the fit of this model with the other two choices of the travel rate.

To quantify the uncertainty of the estimated growth exponent, we performed a Bayesian analysis for this statistical model with diffuse priors. The prior we used for *r* is an exponential distribution with mean log(2) /7.4, matching the previously reported doubling time using cases in Wuhan. The prior we used for *WI*_0_ is such that the number of new infections on January 1, 2020 has prior mean 50 and standard deviation 100. We used an exponential prior for *OR* whose mean is computed from dividing the estimated number of travels from Wuhan to the selected regions by the population of Wuhan. We expected that the posterior of the number of new infections *WI*_*t*_ would be very sensitive to the choice of the prior mean of *OR*, but the posterior of the growth exponent would be insensitive to the choice of prior.

We fitted the models for 100 realizations of the infection counts from January 1 to January 20, expecting the public confirmation of human-to-human transmission on January 20 might have lowered the epidemic growth in the next few days. We did not use infections before January 1 because the simulation could be unreliable for early infections. We also fitted the models using all infections as a sensitivity analysis. The Bayesian models were fitted using the *rstan* software package in *R* (Carpenter et al., 2017). We computed the posterior means and credible intervals by pooling posterior samples for different realizations of the infection counts.

## Results

### Simulated infection time

We obtained the distribution of the mean counts of new infections among the cases and found that it was growing exponentially till January 18 but was dropping from January 19 to 23 (Figure 2, Panel A). This qualitative behavior contradicts an exponential growth model for *OI*_*t*_ (third choice for *OR*_*s*_). However, the correction factor *N* − *t* + 1 is our working model predicts the mean counts to be decreasing when

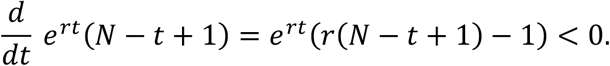

**Figure 2.**
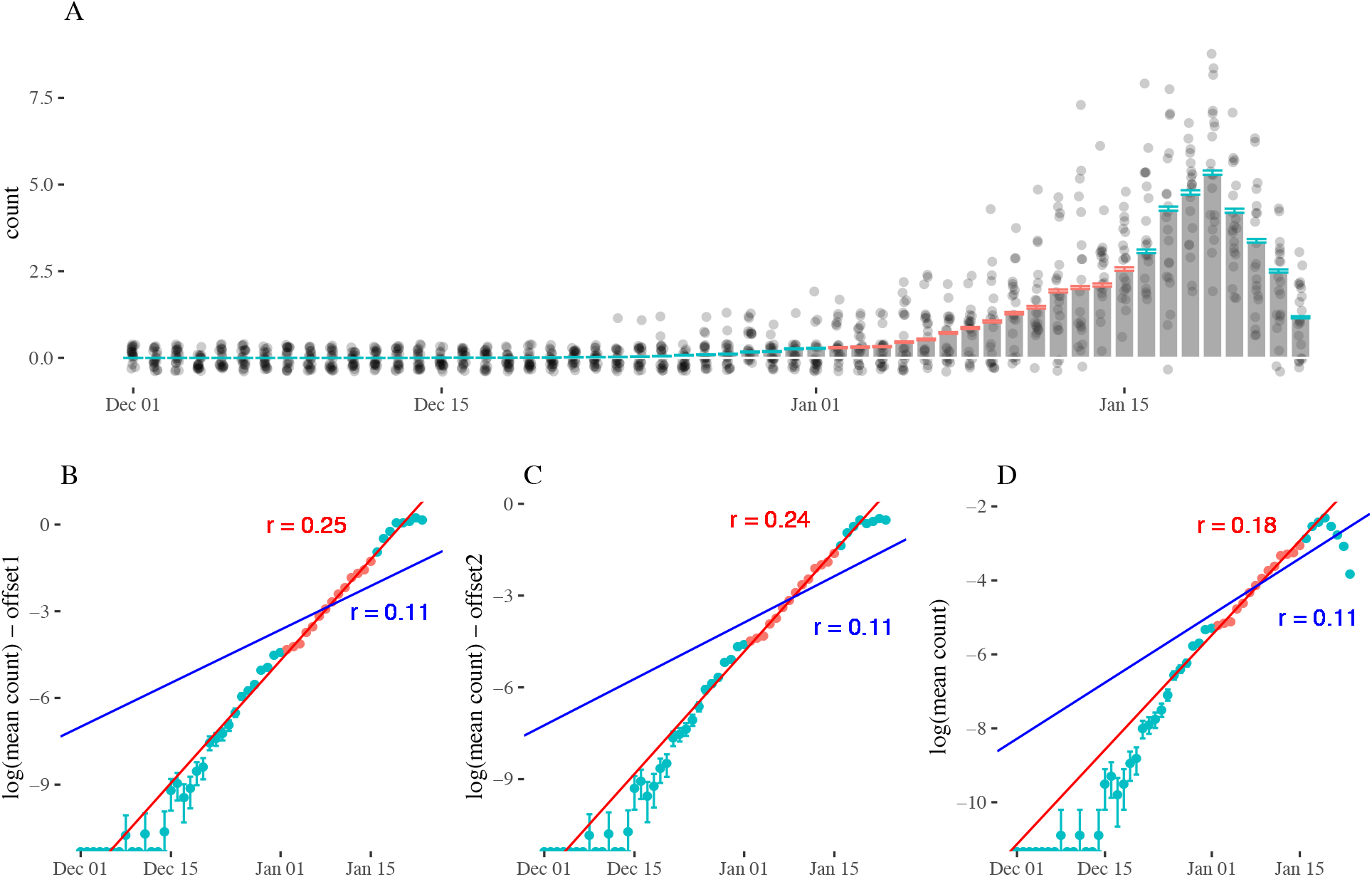
Fitted models using mean counts of new infections. Panel A: Grey bars are the mean counts of new infections based on infection dates simulated from symptom onset dates and assumed incubation period that are consistent with the patients’ exposure history to Wuhan. Red and blue error bars are the one standard error of the mean counts due to Monte-Carlo simulations. Grey dots are 10 realizations of the simulated infection counts. Panel B,C&D: Fitted linear regression for logarithm of the mean counts versus time with different offsets (B: assume constant travel rate; C: assume travel rate doubled from January 21 to January 23; D: ignore the travel ban out of Wuhan since January 23, 2020). Red lines are linear regressions fitted using data in panel A from January 1 to January 15. Blue lines shows growth exponent r=0.11 which correspond to the estimated doubling time of 6.4 days in (Wu et al., 2020).

This equation shows that the stationary point is 1/*r* days before January 24. The empirical stationary point between January 19 and 20 puts *r* between 0.2 and 0.25. Error bars in Panel A also show that the uncertainty of the estimated mean infection counts due to using Monte-Carlo simulations is small.

In fitted linear regressions for logarithm of mean infection counts versus, Modeling the effect of the travel ban on January 23 (Panels B and C, Figure 2) visually improves the fit to the data when compared to ignoring the travel ban (Panel D, Figure 2). The fitted growth exponent using our working model was *r* = 0.25 (Panel B). This is consistent with our prediction of *r* in the last paragraph based on the decreasing infections after January 19. On the contrary, if we assumed the travel rate doubled in the last three days, a similar calculation shows that the stationary point in Panel A should be around January 16. These qualitative assessments show that our working model provide superior fit to the data.

To aide comparison, we included lines in panels B, C, and D, in Figure 2 corresponding to a growth rate of *r* = 0.11 or a doubling time of 6.4 days reported by a previous analysis (Wu et al., 2020). This estimate did not fit the average infection counts well.

### Epidemic growth

In our Bayesian model fitted using infections before January 21 and assuming constant travel rate, the posterior mean of the growth exponent *r* was 0.25 (95% CrI, 0.17 – 0.34). This corresponds to a doubling time of 2.9 days (95% CrI, 2.0 days – 4.1 days) and basic reproduction number of 5.7 (95% CrI, 3.4 – 9.2). The basic reproduction number is computed using the serial interval previously reported for 2019-nCoV (Li et al., 2020).

We further performed a thorough sensitivity analysis of our results (Table 1 and Online Supplement) using a different estimation period, the other two travel rates, and the incubation period and serial interval reported for the severe acute respiratory syndrome (SARS) (Donnelly et al., 2003; Lipsitch et al., 2003). We found that our estimated epidemiological parameters are relatively insensitive to these choices unless we ignore the outbound travel ban on January 23. We also performed a sensitivity analysis to the prior chosen for the rate of travel. Not surprisingly, we found that the estimated growth exponent is not sensitive to the choice, but the total number of infections are very sensitive to the choice (Online Supplement).

**Table 1.**
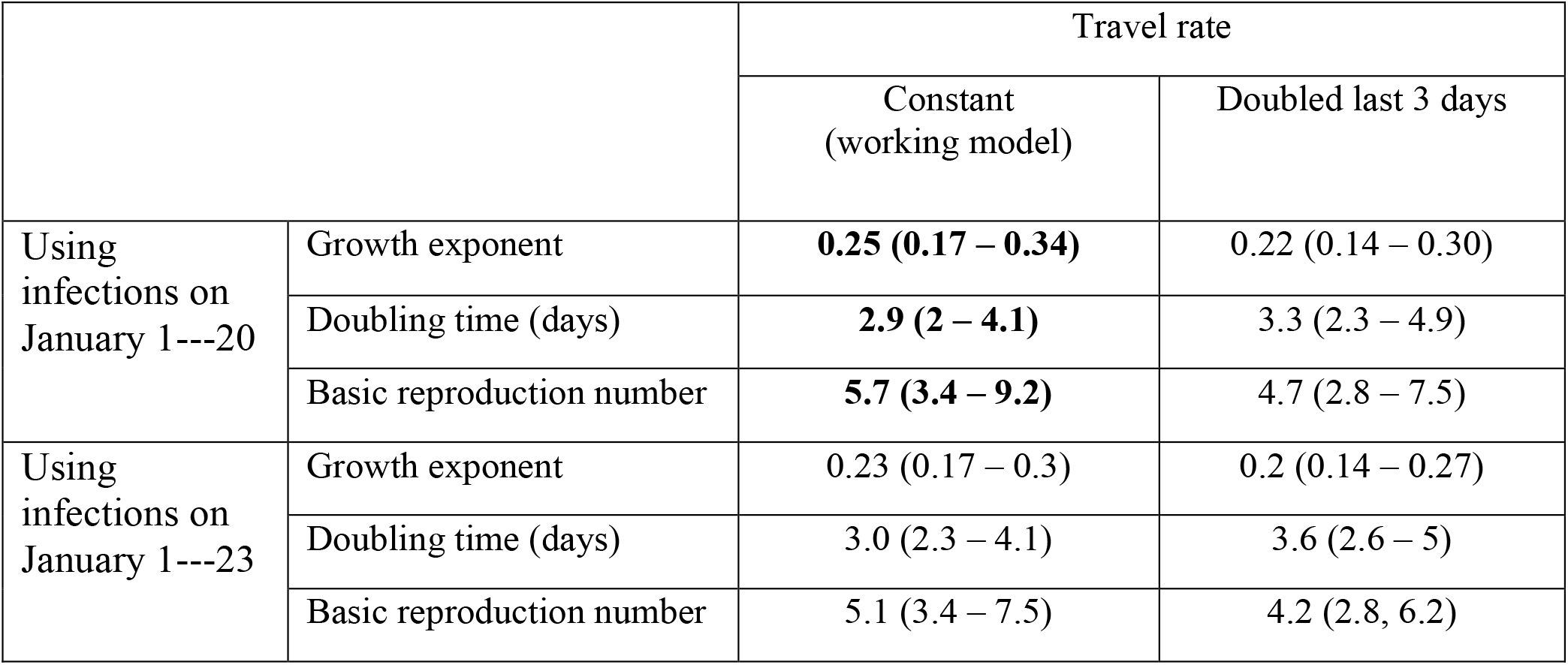
Estimated parameters for epidemic growth rate using different periods of infection and different assumptions on the travel rate. Estimated parameters are in the form of “posterior mean (95% credible interval)”.

|  |  | Travel rate |  |
| --- | --- | --- | --- |
|  |  | Constant<br>(working model) | Doubled last 3 days |
| Using<br>infections on<br>January 1---20 | Growth exponent | **0.25 (0.17 – 0.34)** | 0.22 (0.14 – 0.30) |
|  | Doubling time (days) | **2.9 (2 – 4.1)** | 3.3 (2.3 – 4.9) |
|  | Basic reproduction number | **5.7 (3.4 – 9.2)** | 4.7 (2.8 – 7.5) |
| Using<br>infections on<br>January 1---23 | Growth exponent | 0.23 (0.17 – 0.3) | 0.2 (0.14 – 0.27) |
|  | Doubling time (days) | 3.0 (2.3 – 4.1) | 3.6 (2.6 – 5) |
|  | Basic reproduction number | 5.1 (3.4 – 7.5) | 4.2 (2.8, 6.2) |

## Discussion

Our analysis reported a much higher growth rate of the early 2019-nCoV outbreak in Wuhan. A direct comparison can be drawn with a previous analysis that also used internationally confirmed cases to infer the epidemic size and growth in Wuhan (Wu et al., 2020). However, their estimated doubling time is 6.4 days (95% CrI, 5.8 days – 7.1 days). Another report (Li et al., 2020) using the cases in Wuhan estimated the doubling time to be 7.4 days (4.2 days – 14 days). These estimates did not fit our simulated distribution of infections well, even with our impossibly extreme assumption of the travel rate (Figure 2, Panel D). This shows that the epidemic may have been spreading much faster in Wuhan than these earlier estimates.

There are several possible explanations for the large difference in epidemiological estimates. One possible reason is the different study sample. The previous reports used internationally confirmed cases as of January 28 and the first 425 confirmed cases in Wuhan. In comparison, we used confirmed cases in 6 selected countries and regions with detailed case reports as of February 4. Our selected countries and regions also have good public health surveillance systems and could more easily identified suspected cases in border control. Another key distinction is that our analysis used the simulated infection time instead of the noisier symptom onset as in the previous analyses. By using the cases’ travel history (in particular, international arrival date), we can narrow down when the cases were infected. This reduces the noise in our estimates.

A third distinction with the analysis by (Wu et al., 2020) is that our model takes into account the restricted outbound travel from Wuhan since January 23, 2020 --- a milestone event in this epidemic. The analysis by (Wu et al., 2020) used a more complicated susceptible-exposed-infectious-recovered (SEIR) model and only accounted for the quarantine of Wuhan in their forecast but not in their estimation. Ignoring the travel ban since January 23 could also produce much smaller growth rate in our model. However, we have convincingly demonstrated that this leads to very poor fit of the distribution of infection time.

Our analysis should be viewed in terms of its limitations. The international cases are only “shadows” of the epidemic in Wuhan and we relied on the assumption that they form a representative sample. We used a simple exponential growth model for the new infections and did not account for the dynamics of the epidemics like a SEIR model. We assumed a constant rate of travel in the study period which might not approximate the travel pattern in Wuhan---a transportation hub in central China---very well. In particular, millions of people (substantial proportion of the Wuhan population) traveled back home from Wuhan before the Lunar New Year, which we did not consider in our model. Finally, our estimates are relatively but not entirely insensitive to the incubation period which are crucial in simulating the infection time.

Despite these potential limitations, we convincingly demonstrated that a simple theoretical model --- exponential growth with correction for the travel ban on January 23 --- provides very good fit to the internationally confirmed cases with detailed case trajectories. Our results suggest that the early outbreak of 2019-nCoV could have been spreading much faster in Wuhan than previous estimates. This has important implications in designing prevention measures to control the outbreak in other cities in China and around the world. On the more positive side, our simulated infection rates in January 21 to 23 showed visually significant departure from the previous growth pattern (Panel B, Figure 2). This gives some evidence that the prevention measures following the public confirmation of human-to-human transmission in the evening of January 20 were effective.

## Data Availability

Data for this study are from public sources. We have kept the data and computer programs used in our analysis at the link below, so the results reported in this article and previous versions of our analysis are fully reproducible.

https://github.com/qingyuanzhao/2019-nCov-Data

## Data and reproducibility

Data for this study are from public sources. We have kept the data and computer programs used in our analysis at https://github.com/qingyuanzhao/2019-nCov-Data, so the results reported in this article and previous versions of our analysis are fully reproducible.

## Acknowledgement

The authors have no conflict of interest to declare. We thank Michael Levy for providing helpful comments on this analysis.

